# ALFIE: Anatomy-aware enhancement of Low FIEld 64mT T2-weighted neonatal brain MRI for structural analysis

**DOI:** 10.64898/2026.08.25.26361317

**Authors:** Paul Cawley, Alena Uus, Kathleen Colford, Francesco Padormo, Rui Teixeira, Ines Tomazinho, UNITY Consortium, Steven C.R. Williams, A. David Edwards, Jonathan O’Muircheartaigh, Tomoki Arichi, Joseph V. Hajnal, Mary A. Rutherford

**Affiliations:** Research Department of Early Life, Imaging, School of Biomedical Engineering, and Imaging Sciences, King’s College, London, London, UK; Neonatal Intensive Care Unit, Evelina, Children’s Hospital London, St Thomas’ Hospital, London, UK; MRC Centre for Neurodevelopmental, Disorders, King’s College London, London, UK; Hyperfine Inc. Guilford, CT, USA; Department of Neuroimaging, Institute of Psychiatry, Psychology and Neuroscience, Kings College London, London, United Kingdom; Department of Forensic and Neurodevelopmental Science, Institute of Psychiatry, Psychology and Neuroscience, Kings College London, London, United Kingdom; Research Department of Imaging Physics and Engineering, School of Biomedical, Engineering and Imaging Sciences, King’s, College London, London, UK

**Author notes:** P.C. and A.U. are joint first authors. **Correspondence** Alena Uus, King’s College London, London, UK.

**Keywords:** Neonatal brain, low field MRI, image enhancement, segmentation

## Abstract

**Purpose:** To develop and evaluate an anatomy-aware deep learning framework for enhancement of neonatal 64mT T2-weighted MRI that improves anatomical visibility while preserving native ultra-low-field contrast and enabling quantitative structural analysis.

**Methods:** A multitask network, jointly performing image enhancement and tissue segmentation, was trained on 75 and evaluated on 20 paired neonatal 64mT/3T MRI datasets spanning a broad range of gestational ages and pathologies. To preserve native 64mT contrast, 3T images were locally harmonized before training. The framework also generated quality-control maps and regional volumetric measurements. Volumetric agreement was further assessed in 40 paired term-born control datasets.

**Results:** Enhanced 64mT images showed improved image quality metrics and better delineation of cortical, deep gray matter, ventricular, white matter, and posterior fossa structures while maintaining native contrast characteristics. Tissue segmentations demonstrated good agreement with reference 3T labels. Volumetric measurements showed excellent correspondence with 3T across major tissue compartments, with only small systematic regional biases.

**Conclusions:** Anatomy-aware enhancement enables automated tissue segmentation and volumetric analysis directly from neonatal 64mT MRI while preserving native image contrast. These findings support the feasibility of quantitative neonatal neuroimaging at ultra-low field.

## Introduction

Portable ultra-low-field MRI systems such as the 64mT Swoop®scanner (Hyperfine, Inc., Guilford, USA) have emerged as a promising approach for expanding access to neonatal neuroimaging [1, 2]. Unlike conventional 1.5T and 3T scanners, portable systems enable bedside imaging, reducing the need to transport critically ill infants and increasing accessibility in intensive care units and low-resource settings. Recent neonatal-specific acquisition protocols have demonstrated that 64mT MRI can visualize major neuroanatomical structures and clinically relevant abnormalities [3, 4]. Additional studies have shown the feasibility of point-of-care neonatal imaging during natural sleep [5, 6], improved diagnostic performance compared with cranial ultrasound [7], and reliable assessment of conventional ultrasound biometry [8].

Despite these advances, widespread adoption of neonatal 64mT MRI remains limited by the intrinsic constraints of low-field imaging. Compared with conventional scanners, 64mT images exhibit reduced signal-to-noise ratio (SNR), lower spatial resolution, increased partial-volume effects, and altered tissue contrast. These differences arise from both field-dependent relaxation properties, including shorter T1 values and reduced susceptibility effects, and differences in acquisition and reconstruction protocols [3, 9]. Consequently, relative tissue signal intensities differ from those observed at 3T. E.g., an example in Fig. 1 illustrates that deep gray matter and posterior fossa structures tend to exhibit lower relative signal intensity at 64mT, whereas white matter demonstrates a broader intensity distribution.

**FIGURE 1.**
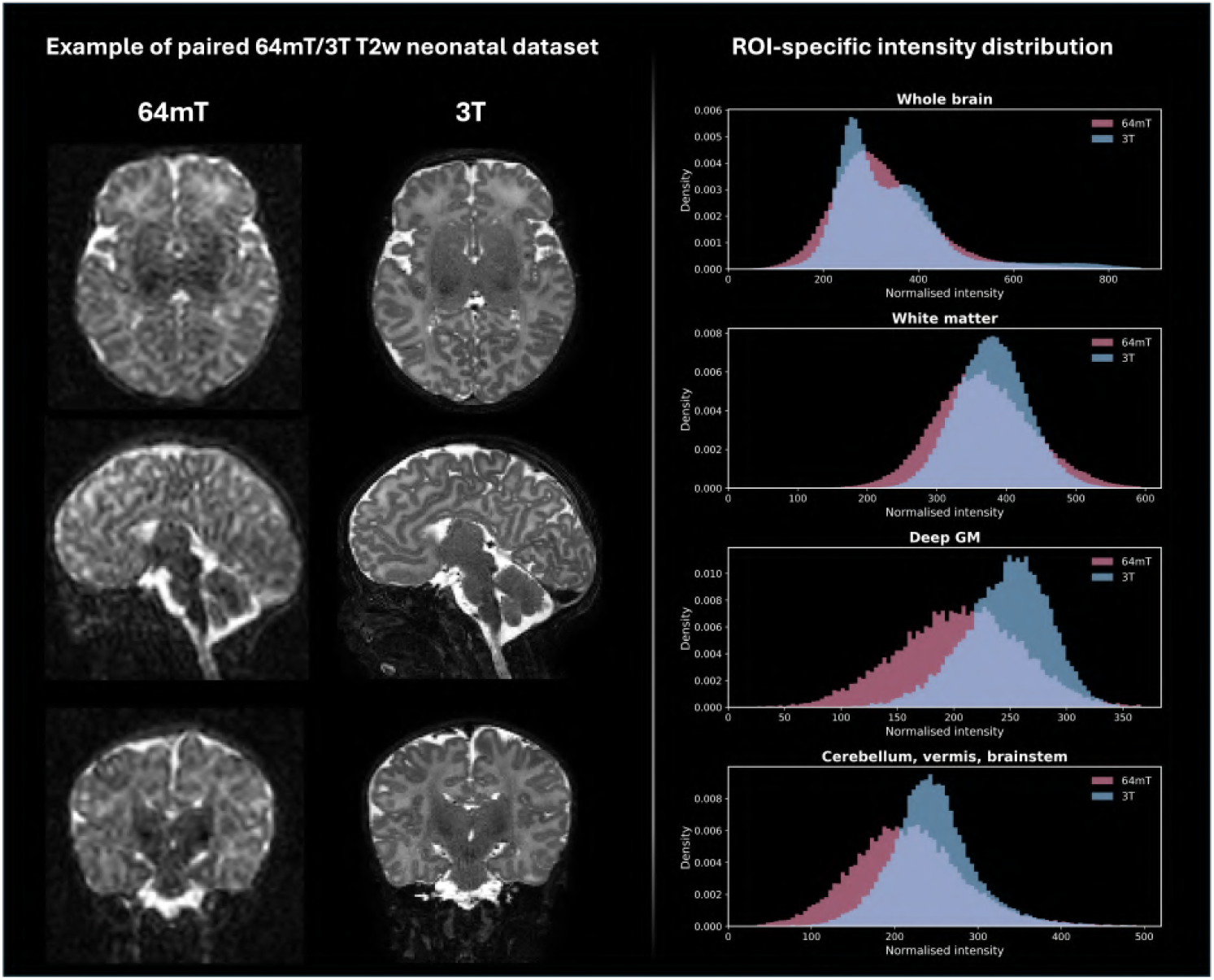
Example of same day paired 64mT / 3T neonatal dataset acquired using optimised low field protocol [3]. The intensity histograms were created from aligned and normalized 64mT and 3T images within the same tissue class segmentation regions.

In terms of both radiological assessment and downstream analysis applications at 64mT, low SNR and resolution reduce the visibility of tissue boundaries and finer anatomical features thus limiting confidence in visual image interpretation as well as delineation of structures for volumetric analysis. The challenge is particularly pronounced in neonatal brain imaging with small, rapidly developing structures and dynamic changes in tissue properties.

Recent studies explored various deep learning (DL) solutions for low-field MRI enhancement for adult and pediatric populations, including image synthesis, super-resolution, and generation of high-field-like images from low-field acquisitions [10, 11, 12, 13, 14]. Similar approaches have also been applied to neonatal MRI, demonstrating the feasibility of synthesizing 1.5T- and 3T-equivalent T2w images from 64mT scans using generative adversarial networks and diffusion models [15, 16, 17]. Yet, while these contrast transfer methods can improve visual image quality, they are primarily designed to reproduce the appearance of conventional high-field MRI. However, since contrast differences between 64mT and 1.5/3T MRI arise from field-dependent relaxation properties and acquisition protocols, synthetic contrast mapping may alter physically meaningful tissue characteristics. This is especially critical for interpretation of abnormal cases (e.g, hypoxic-ischemic encephalopathy) where radiological diagnosis relies on subtle intensity patterns ([18, 19]. Thus, neonatal 64mT images are not simply degraded high-field MRI, but have native contrast information. Therefore, a powerful approach could be to deploy image enhancement to improve anatomical visibility while preserving native 64mT contrast and biologically meaningful tissue relationships. Furthermore, generative models are also susceptible to uncertainty and hallucinated features, potentially compromising the reliability of synthetic images [20, 21]. In terms of structural analysis, although several segmentation frameworks have been reported for paediatric and infant 64 mT MRI [22], no dedicated tools currently exist for the segmentation of low-field neonatal MRI. And although recent DL tissue segmentation pipelines for conventional neonatal MRI support volumetric analysis and normative modelling [23], they are not directly applicable to 64mT.

### Contributions

This work addresses the existing challenges by leveraging the advanced neonatal 64mT MRI acquisition protocol [3] with a cohort of paired 64mT/3T datasets to develop an anatomy-aware DL enhancement framework for neonatal 64mT T2w MRI. The proposed pipeline combines local intensity harmonization with multitask deep learning, jointly optimizing image enhancement and tissue segmentation. Rather than replicating high-field image appearance, the framework aims to improve delineation of neonatal brain structures while preserving anatomically meaningful 64mT contrast characteristics. The enhanced images are subsequently integrated with regional volumetry measurements, enabling quantitative structural analysis of neonatal 64mT MRI. In addition, segmentation confidence and model uncertainty estimates are incorporated for quality control and identification of potentially unreliable regions.

## Methods

### Cohort and datasets

A total of 115 paired neonatal MRI datasets were acquired at St Thomas’ Hospital, London, under UK NHS Research Ethics approvals 12/LO/1247 and 19/LO/1384. The cohort (Fig. 2) comprised infants spanning a wide range of gestational age at birth (23.29 – 42.14 weeks) and PMA at scan (28.71 - 51.57 weeks) and included cases with normal MRI findings, minor abnormalities (e.g., punctate white matter lesions, cysts, and mild ventriculomegaly), acquired brain injury (including hypoxic-ischemic encephalopathy, infarction, hemorrhage, and white matter injury), congenital brain dysgenesis, and combined congenital malformations with superimposed acquired injury.

**FIGURE 2.**
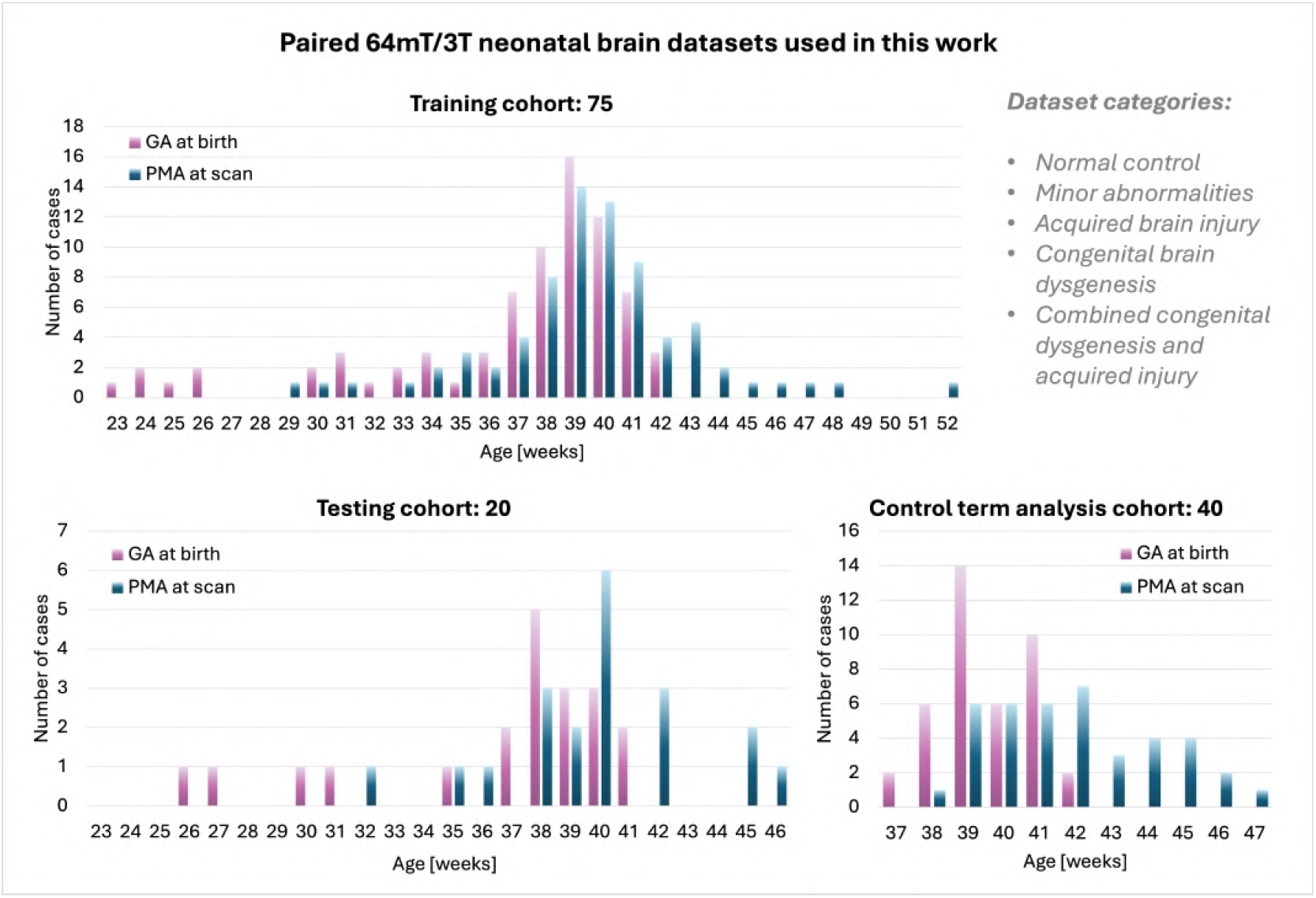
Paired 64mT/3T neonatal MRI datasets used in this work.

3T MRI data were acquired on a Philips Achieva system (Philips Healthcare, Best, The Netherlands) using a neonatal T2-weighted turbo spin-echo protocol with the following parameters: TR = 12,000 ms, TE = 156 ms, in-plane resolution = 0.8 × 0.8 mm, slice thickness = 1.6 mm. For each subject, the highest-quality T2w image stack was selected for analysis.

64mT MRI data were acquired using a portable Swoop® low field scanner (Hyperfine Inc., Guilford, CT, USA) with an optimized neonatal T2w protocol described by Cawley et al. [3]. Imaging was performed using a 3D fast spin-echo sequence with TR = 2000 ms, TE = 281–303 ms, and isotropic voxel size of 1.75–2.00 mm. For each subject, the highest-quality T2w stack was selected for analysis.

The primary inclusion criteria were the availability of acceptable quality T2w images at both field strengths, including absence of visible motion artifacts or severe signal loss, full brain coverage, and visibility of major anatomical structures, and a time interval of less than three days between paired 64mT and 3T acquisitions. The training (75) and testing (20) subsets include term and preterm subjects at wide range of anomalies. The additional cohort selected for segmentation analysis (40) includes only term born control subjects and it was comprised of the datasets used on training and testing as well as an additional 20 independent datasets.

As preparation for training and testing, 3T images underwent automated brain extraction and reorientation to standard radiological space [23]. Subsequently, the paired 64mT images were aligned to the corresponding 3T images using MIRTK affine and global non-linear free-form deformation [24] registration with local normalized cross-correlation similarity metric. The 3T brain masks were also propagated to the native 64mT space.

Next, multi-tissue brain segmentation of 3T images was performed using the Multi-BOUNTI pipeline [23]. All segmentations were visually inspected and manually refined when necessary, primarily in subjects with severe structural abnormalities. For the purpose of this work, lobe-specific (for cortical gray, deep gray and white mater) ROIs were combined into global left/right (L/R) hemisphere specific labels, resulting in 15 labels in total.

### Proposed image enhancement pipeline

An overview of the proposed framework is presented in Fig. 3. In summary, the aim is to enhance 64mT images in terms of the anatomical feature definitions while preserving the native contrast and provide simultaneous tissue parcellation output. This is achieved via spatial standardization followed by an image enhancement step via a multi-channel network that learns contrast mapping from 64mT T2w images to 3T T2w images with locally matched intensity distribution and ensures anatomy awareness via global tissue ROI segmentation.

**FIGURE 3.**
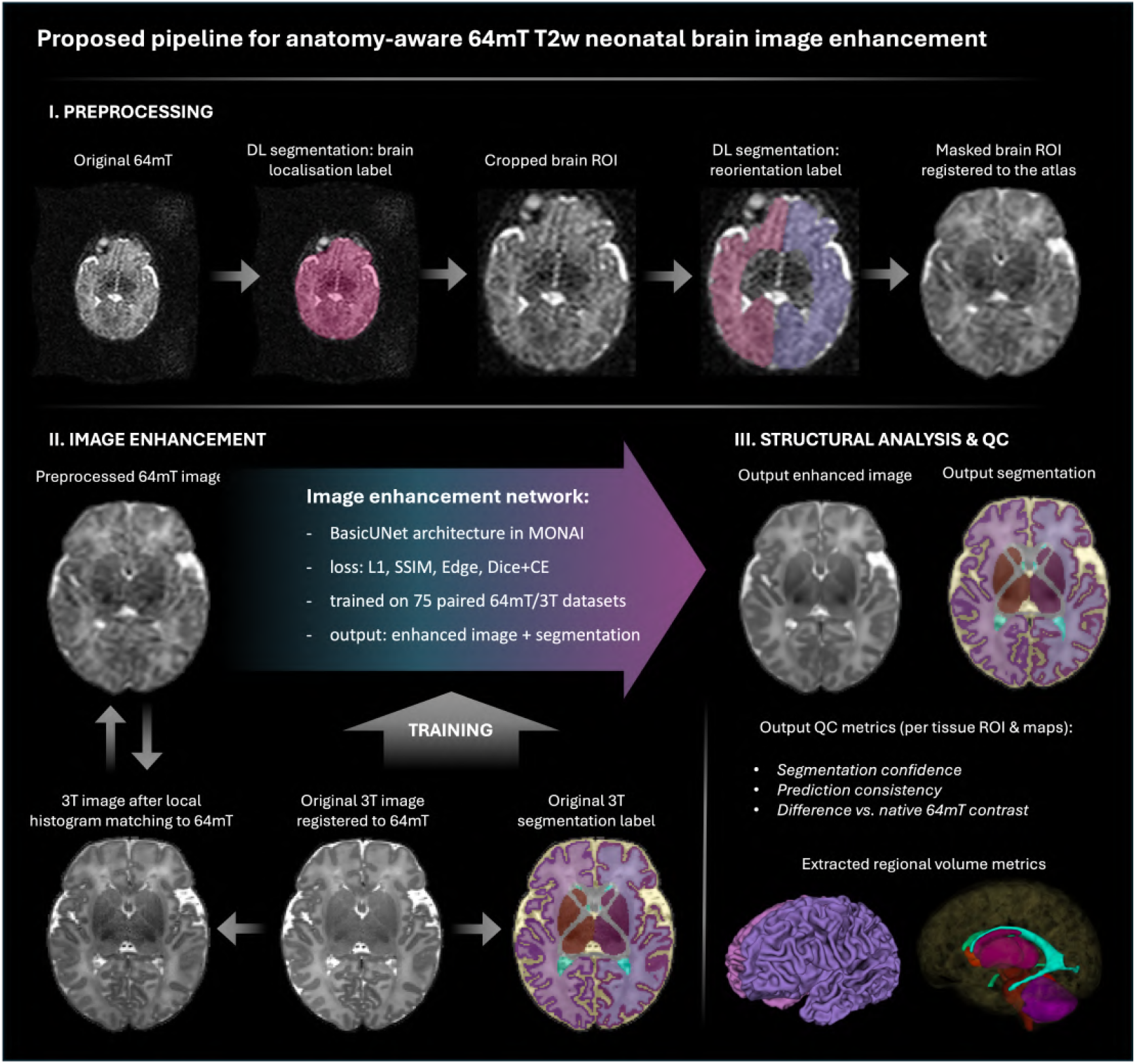
Overview of the proposed pipeline for anatomy-aware contrast enhancement for 64mT neontal brain MRI.

### Preprocessing

Similarly to the preprocessing steps in [23], at first, the brain is globally localized in raw 64mT image. Next, the cropped brain is segmented to provide L/R hemisphere ROIs that will be used for global affine registration to the standard radiological space. The brain localisation and L/R reorientation ROIs networks are based on classical 3D Attention UNet [25] architecture in MONAI [26] each trained on 75 64mT datasets with standard augmentations. After registration, the transformed 64mT image is then masked and resampled with zero-padding to 128 x 128 x 128 grid and along with the brain mask passed to the next step.

### Anatomy-aware image enhancement network

#### Local intensity harmonization

At first, for training of the 64mT image enhancement network we needed to account for regional intensity variations and spatially varying contrast differences between 3T and 64mT while preserving the native low field contrast domain.

This was achieved by using sliding-window local histogram matching to make 3T images have 64mT like tissue contrast distribution. All 3T images were matched to the corresponding 64mT images using overlapping three-dimensional patches (block size 20, stride 10 voxels). Matched patches were subsequently combined using weighted averaging to generate spatially consistent harmonized images.

#### Network architecture

An image enhancement step was implemented as a multitask 3D convolutional neural network implemented in PyTorch and MONAI [26]. The network was based on the MONAI BasicUNet architecture, a fully convolutional encoder-decoder network with skip connections between corresponding resolution levels. The model comprised feature channels of 32, 32, 64, 64, 128, 128, instance normalization, and leaky-ReLU activation functions.

A single-channel harmonized 64mT T2w image was provided as input and processed through a shared encoder-decoder backbone. The network generated 17 output channels comprising one enhanced image channel and 16 segmentation channels. The segmentation outputs corresponded to background, cerebrospinal fluid (CSF), left and right cortical gray matter (GM), left and right white matter (WM), left and right deep gray matter, internal white matter background, left and right ventricles, cavum, left and right cerebellar hemispheres, vermis, and brainstem.

Image enhancement and tissue segmentation were optimized jointly, providing anatomical supervision during training. This multitask formulation encouraged preservation of tissue boundaries and regional anatomy while reducing anatomically implausible image modifications.

#### Network training

The dataset was randomly divided into training (n=72) and validation (n=3) subsets. Network training was performed using paired 64mT and locally harmonized 3T images together with corresponding Multi-BOUNTI tissue segmentations and brain masks. All images were intensity-normalized to the range [0,1].

Training was performed for 40,000 iterations using the AdamW optimizer with learning rate 10^−4^, weight decay 10^−5^ with a batch size of one. Data augmentation included random affine rotations ((±5^◦^) and left-right flipping within MONAI.

#### Loss functions

Network optimisation was performed using a weighted combination of image reconstruction and regional tissue segmentation losses. The image reconstruction loss (*L*_img_) combined masked voxel-wise L1, structural similarity (SSIM), and edge-preservation terms. The L1 term promotes voxel-wise fidelity between the enhanced image and the locally harmonized target image. The SSIM term preserves local structural information and neighborhood-level contrast. The edge-preservation term compares 3D Sobel gradient-magnitude maps of the enhanced and target images, encouraging preservation of tissue boundaries and reducing excessive smoothing. The segmentation branch was optimised using a combined Dice and cross-entropy loss (*L*_seg_). The total loss function was: *L*_total_ = *λ*_*L*1_*L*_*L*1_ + *λ*_*SSI M*_ *L*_*SSI M*_ + *λ*_*edge*_ *L*_*edge*_ + *λ*_seg_*L*_seg_.

Loss weights were selected empirically using validation experiments to balance image fidelity, structural preservation, and anatomical supervision. The final weights were *λ*_*L*1_ = 0.50, *λ*_*SSI M*_ = 0.2, *λ*_*edge*_ = 0.10, and *λ*_seg_ = 0.2. This weighting prioritizes image reconstruction while using segmentation loss as auxiliary anatomical constraints. All image-based losses were computed within the brain mask.

### Quality control and reliability assessment

To support application on previously unseen datasets, the framework incorporates a quality control (QC) module combining segmentation confidence, prediction consistency, and native 64mT contrast-preservation assessment. Segmentation confidence was estimated from voxel-wise softmax probabilities: *C*_*i*_ = max_*c*_ *p*_*i*_ (*c*). Prediction consistency was assessed using left-right flip test-time augmentation and quantified from the difference between enhanced image predictions: 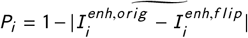, where-· denotes robust normalization to [0, 1]. Native 64mT contrast preservation was evaluated using the input image and corresponding enhanced network output: 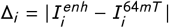.

Voxel-wise confidence, prediction-consistency, and contrast-preservation maps were saved for visual inspection. In addition, ROI-level summary statistics were computed for major tissue regions and exported as .csv files to facilitate identification of anatomical structures potentially affected by enhancement or segmentation uncertainty.

### Pipeline outputs and structural analysis

In order to ensure standard representation for downstream analysis, firstly the original raw 64mT image is rigidly reoriented to the atlas space cropped to the brain ROI and resampled to the 1mm isotropic resolution.

The outputs of the pipeline including enhanced 64mT image, QC metrics, brain mask and multi-tissue segmentation files are then transformed to this reoriented and resampled original image world space. The signal intensity of the enhance image is scaled back to the original 64mT range within the brain ROI mask. The tissue segmentations are then used to compute regional volumetry saved into a .csv file.

### Evaluation

Evaluation was performed on an independent cohort of 20 paired neonatal 64mT/3T datasets not used in training comprising both control and abnormal cases (Fig. 2). Assessment included image fidelity, tissue contrast preservation, segmentation accuracy, structural analysis, expert qualitative review, and ablation experiments. The ablation testing was performed for three different combination of training losses (L1, L1+SSIM+Edge, L1+SSIM+Edge+DiceCE).

Image fidelity was evaluated within the brain mask using peak signal-to-noise ratio (PSNR), structural similarity index measure (SSIM), normalized cross-correlation (NCC), normalized mutual information (NMI), and mean absolute error (MAE) between enhanced 64mT images and the corresponding locally harmonized 3T reference images. Regional contrast preservation was assessed using histogram intersection with the original 64 mT images, where 1 indicates complete overlap. Segmentation performance was assessed using Dice similarity coefficient and absolute percentage volume difference relative to the reference 3T segmentations.

Qualitative assessment was performed independently by a neuroradiologist, a neonatologist, a researcher, and a radiographer with extensive experience in neonatal MRI. Major anatomical structures were scored in terms of improvement of enhanced 64mT vs. native 64mT using a four-point scale (i.e., the native 64mT images have default score 1). The corresponding 3T images were used as an additional reference. Tissue segmentations were scored based on accuracy of anatomical delineations. For image enhancement the grading scheme was: 1 = no improvement; 2 = moderate improvements, presence of uncertainties; 3 = good improvements, presence of minor uncertainties; 4 = significant, close to 3T image quality. For segmentation the grading scheme was: 1 = failed; 2 = moderate quality, presence of inconsistencies; 3 = good, presence of minor inconsistencies; 4 = excellent.

In addition, to assess feasiblity for downstream structural analysis, the complete pipeline was applied to an independent cohort of 40 control term-born infants with 38.43 – 46.71 weeks PMA at scan. Regional tissue volumes derived from enhanced 64mT images were compared with the volumes derived from Multi-BOUNTI segmentations of the corresponding paired 3T images.

### Implementation details

The DL components were implemented in PyTorch (https://github.com/pytorch/pytorch) and MONAI [26]. The image processing steps were implemented using Python and MIRTK library (https://github.com/BioMedIA/MIRTK). The code for training and inference is available at https://github.com/SVRTK/neonatal-low-field-mri-analysis. The full inference pipeline (preprocessing, enhancement, QC, structural analysis) can be executed as a standalone Docker application. The full execution time is less that 10 minutes per case on CPU and less than 5 minutes on GPU versions.

## Results

### Quantitative evaluation

The results of quantitative evaluation for the independent test cohort are summarized in Fig. 4. On average, the enhanced 64mT images showed improved agreement with the corresponding histogram-matched 3T reference images, with consistently higher PSNR, SSIM, NCC, and NMI and lower MAE compared with the original 64mT images. Among the evaluated loss configurations, the multitask model combining image reconstruction, structural similarity, edge-preservation, and segmentation supervision (*L1 + SSIM + Edge + DiceCE*) achieved the best overall performance and provided visibly improved delineation of anatomical structures. Enhanced 64mT images showed high regional histogram overlap with the original 64 mT images (median, 0.82–0.94), consistently exceeding that of the original 3T images.

**FIGURE 4.**
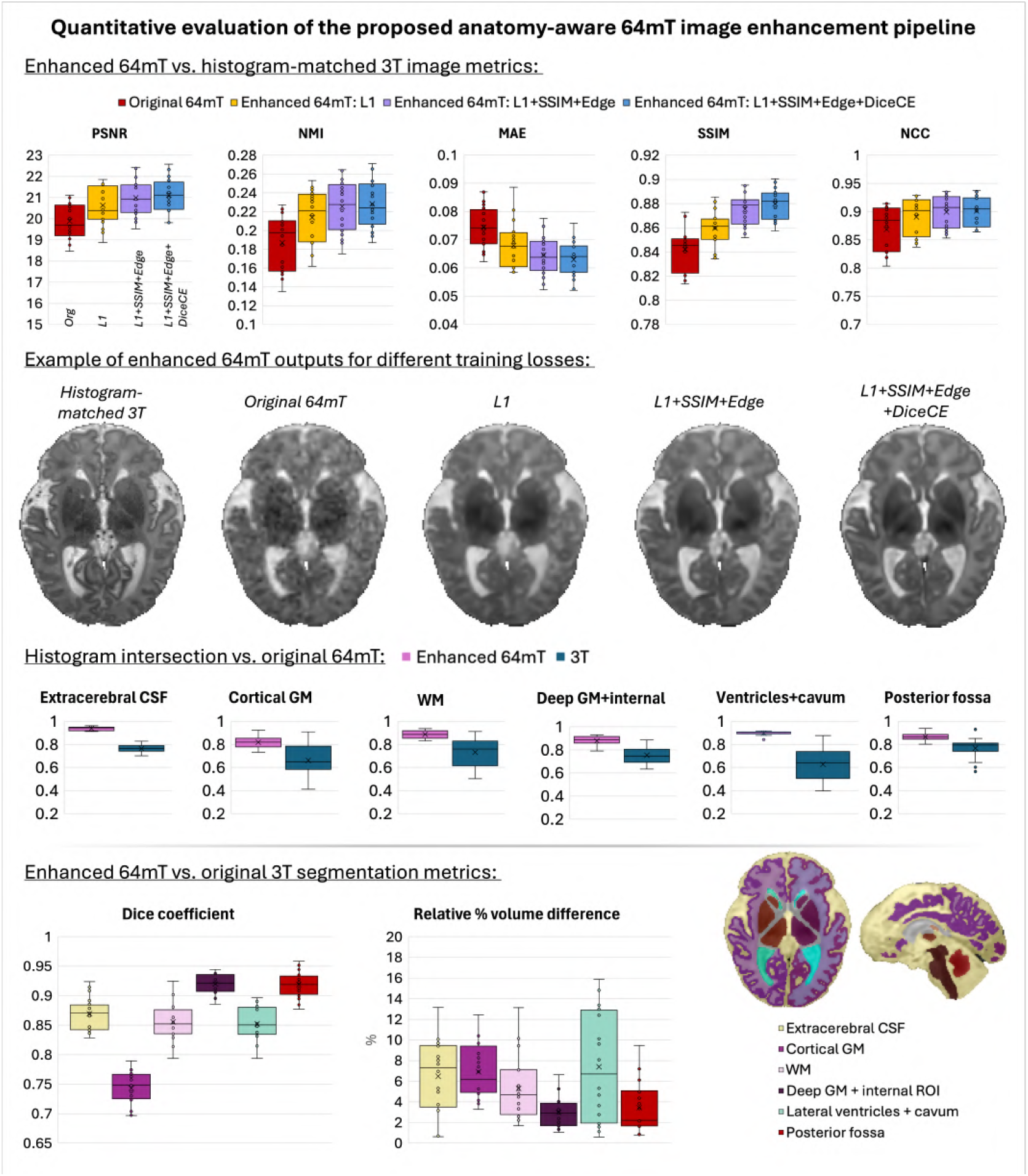
Quantitative evaluation of anatomy-aware enhancement.

Segmentation and volumetric analysis similarly demonstrated high agreement with the reference 3T labels, particularly for deep gray matter and posterior fossa structures. Extracerebral CSF and WM also showed good agreement. Lower Dice values for cortical GM likely reflect both imperfect 64mT-to-3T alignment and the inherently limited cortical visibility at 64mT, resulting in differences in anatomical boundaries between enhanced 64mT and 3T segmentations. The larger variability observed for ventricular measurements was primarily driven by abnormal cases.

Per-case inspection provided additional insight into model behavior. Lower image quality metrics were generally associated with poor native 64mT image quality and reduced visibility of local anatomical features. Segmentation confidence was consistently lower within structural abnormalities (e.g., lesions and ventriculomegaly) and along the cortical ribbon, particularly at later PMA. These regions also exhibited reduced prediction consistency and larger contrast differences, likely reflecting both the limited number of abnormal cases in the training cohort and the heterogeneous nature of neonatal pathology.

### Qualitative evaluation

The results of the qualitative evaluation are summarized in Fig. 5. Overall, reviewers consistently reported improved image quality compared with the native 64mT acquisitions across both control and abnormal cases. The global image enhancement score for the entire brain was 3.25 ± 0.39, indicating substantial improvement while preserving the native 64mT appearance. Among the major tissue classes, the highest scores were achieved for CSF (3.45 ± 0.50) and the lateral ventricles (3.34 ± 0.50), while cortical GM (3.23 ± 0.55), deep GM (3.20 ± 0.51), and WM (3.16 ± 0.50) also demonstrated consistent improvement. Fine anatomical structures important for neonatal diagnosis, including the posterior limb of the internal capsule (PLIC) (3.68 ± 0.45), cerebral aqueduct (3.68 ± 0.50), hippocampus (3.63 ± 0.48), and corpus callosum (CC) (3.24 ± 0.64), showed good enhancement with improved visibility. Although cerebellar anatomy was generally better defined, the posterior fossa received the lowest score (3.09 ± 0.48) due to consistent over-smoothing.

**FIGURE 5.**
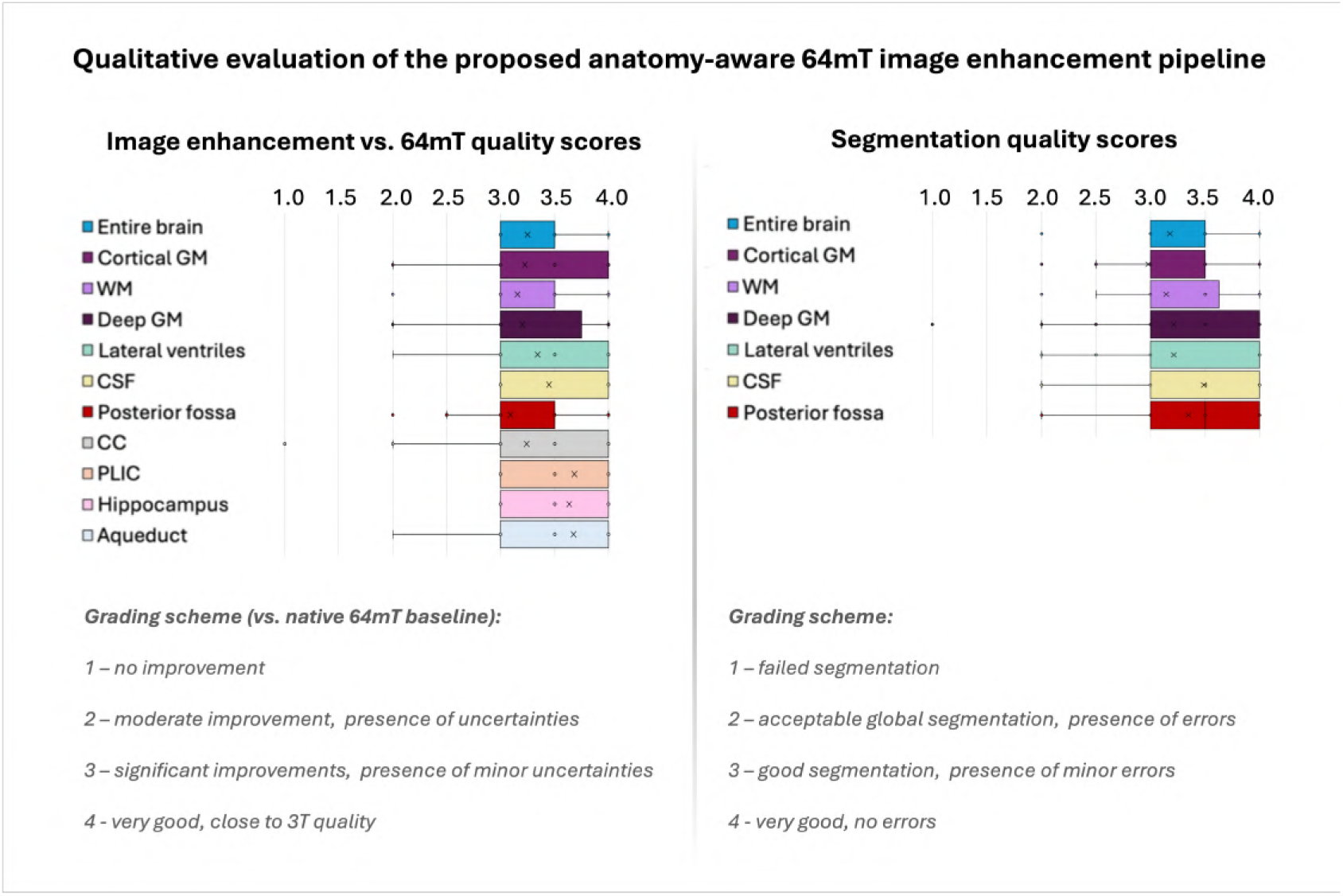
Qualitative evaluation of anatomy-aware enhancement.

**FIGURE 6.**
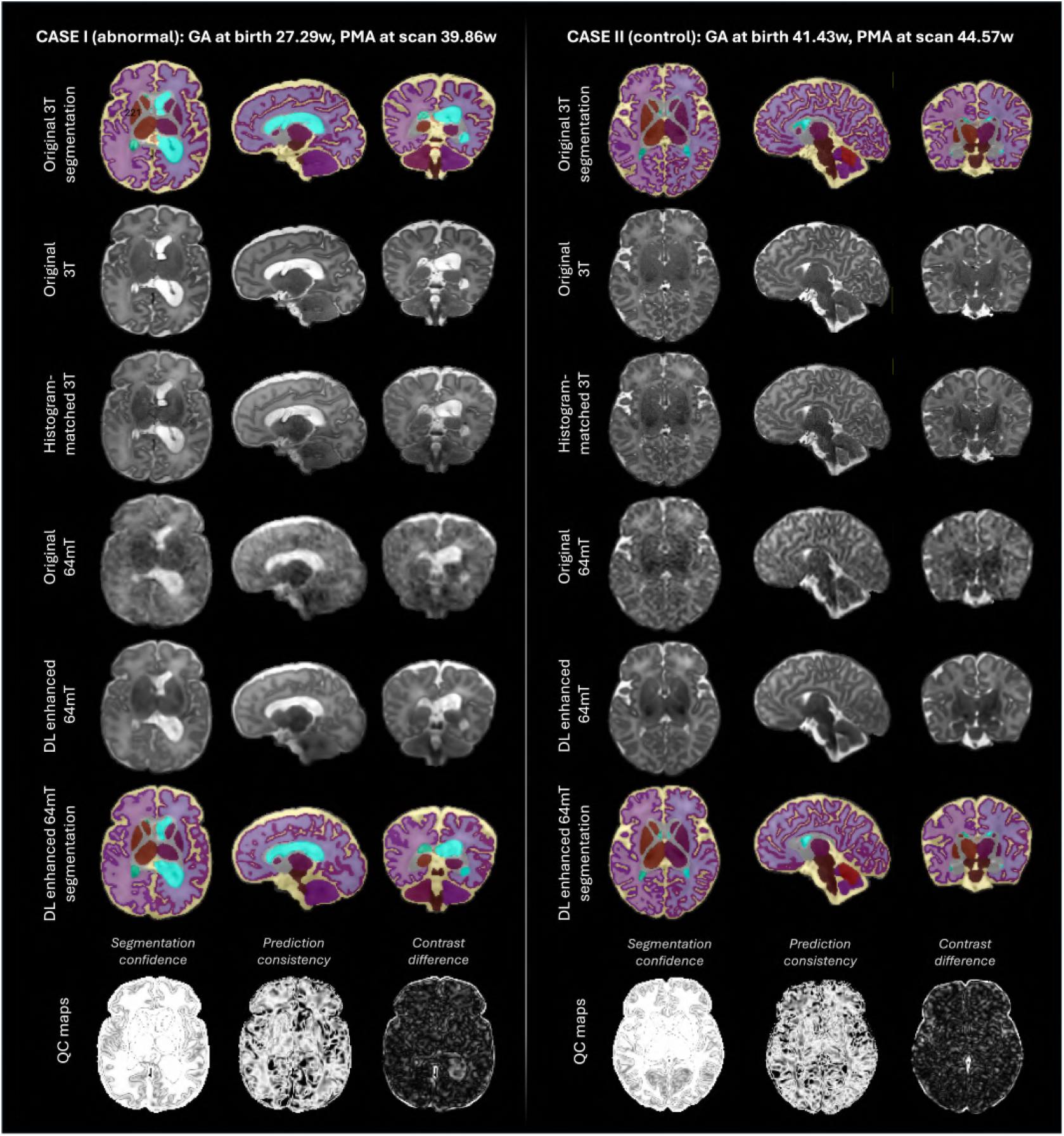
Representative examples from test control and abnormal neonatal datasets showing native 64mT, enhanced 64mT, reference 3T, tissue segmentations and confidence map (Part 1).

Representative examples in Fig. 7-7 demonstrate that the proposed framework improves anatomical visibility while preserving native 64mT contrast characteristics. Visual assessment of the corresponding tissue segmentations similarly indicated good overall quality, with a global whole-brain score of 3.18 ± 0.47. Among the major tissue classes, segmentation scores ranged from 2.98 ± 0.55 for cortical GM to 3.49 ± 0.59 for CSF, with good performance for WM (3.14 ± 0.58), deep GM (3.21 ± 0.67), ventricles (3.21 ± 0.71), and posterior fossa (3.35 ± 0.61). Lower segmentation scores were observed predominantly in subjects with structural abnormalities, particularly focal lesions, where tissue boundaries were altered. Similarly, suboptimal enhancement was mainly associated with severe abnormalities and poor native 64mT image quality, which reduced the visibility of local anatomical features.

**FIGURE 7.**
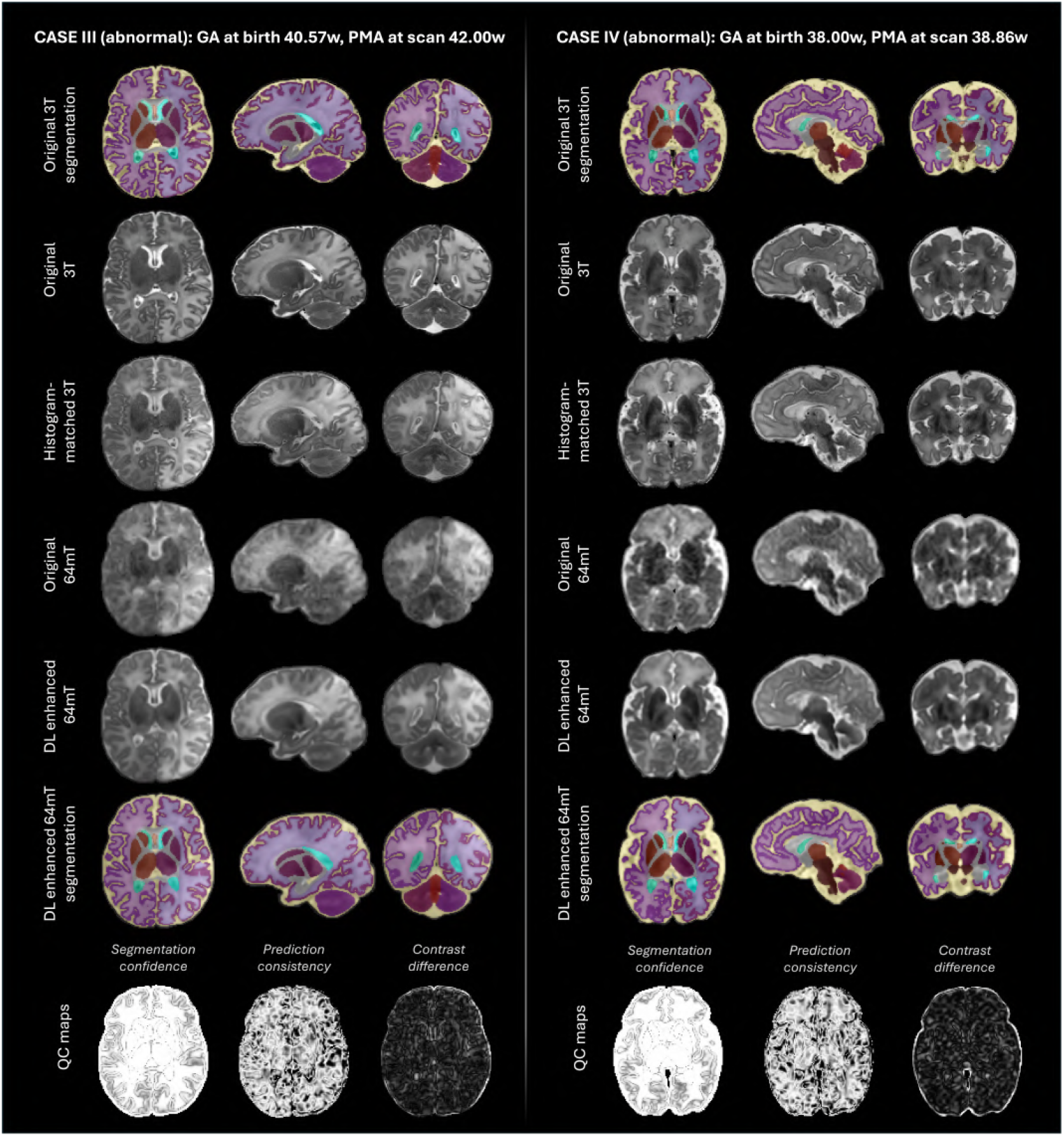
Representative examples from test control and abnormal neonatal datasets showing native 64mT, enhanced 64mT, reference 3T, tissue segmentations and confidence map (Part 2).

### Regional volumetry analysis

The full pipeline was successfully executed for all 40 term 64mT control datasets. All segmentations were visually inspected and considered acceptable for global structural analysis. Corresponding paired 3T scans were segmented independently using Multi-BOUNTI [23].

Fig. 8 summarizes regional volumetric measurements as a function of PMA at scan and compares enhanced 64mT with reference 3T measurements. Regional tissue volumes followed expected maturational trajectories and showed excellent agreement between modalities (*P ear son r* = 0.98–0.997, *ICC* = 0.88–0.99). Although total brain volume was nearly unbiased, systematic differences (*p* < 0.0001) were observed, with slightly larger cortical GM and ventricular volumes (~ 5%) and smaller WM and deep GM volumes at 64mT (4.41%±2.03% on average). These biases likely reflect uncertainty at tissue interfaces and differences in the image resolution (1 mm 64mT vs. 0.5 mm 3T) as well as distortion-related factors. Overall, these findings support the feasibility of large-scale structural analyses at 64mT while highlighting the need for dedicated 64mT normative models.

**FIGURE 8.**
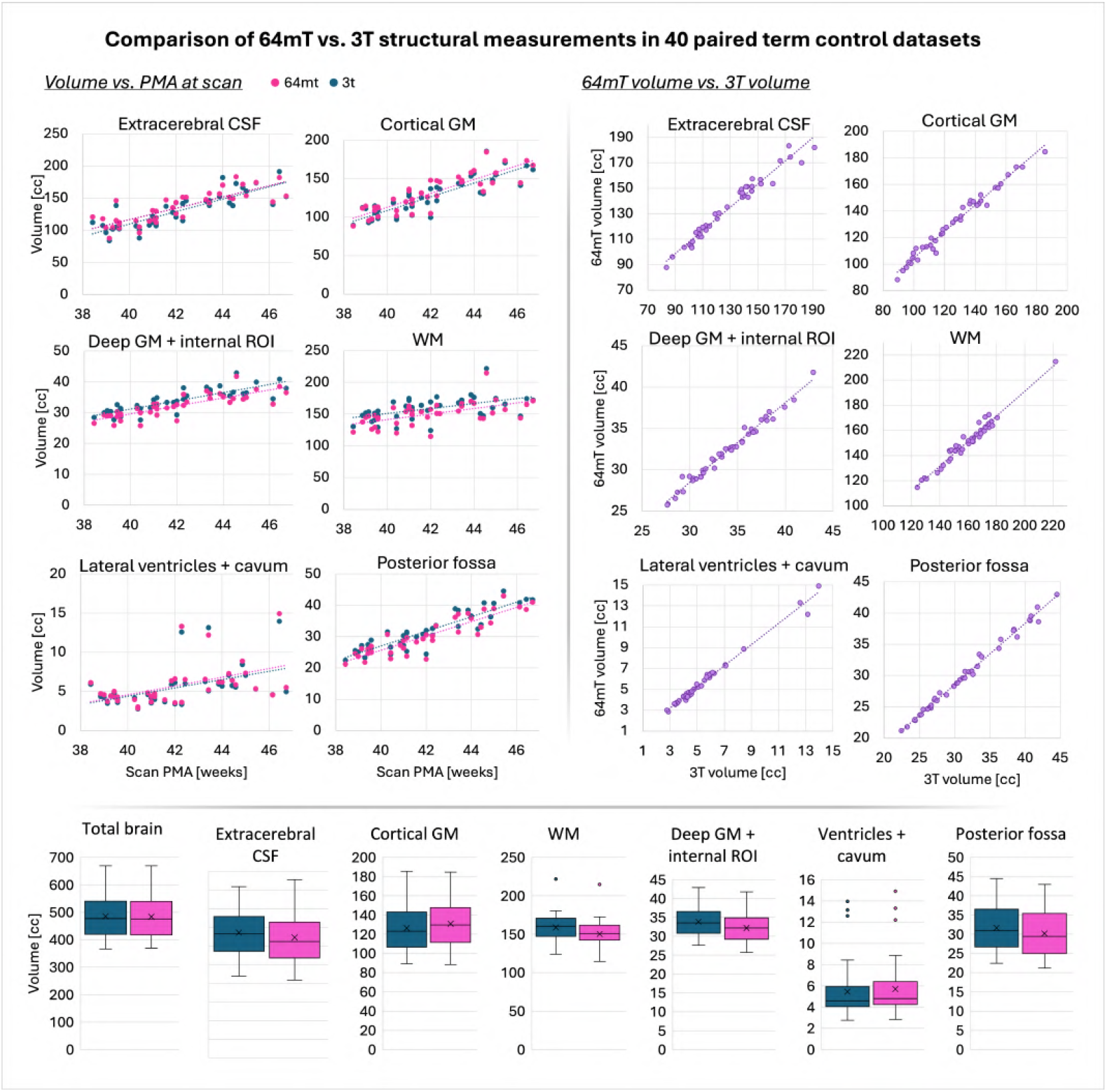
Comparison of 64mT (pink) and 3T (blue) volumetric growth trajectories derived from 40 paired 64mT/3T control term datasets.

## Discussion

Building on the recent advances in neonatal 64mT acquisition [3], this study investigated the feasibility of anatomy-aware enhancement of neonatal 64mT MRI using paired 64mT/3T datasets and a multitask deep learning framework combining image enhancement and tissue segmentation. Unlike previous approaches that aim to synthesize high-field-like images, the proposed method was designed to preserve native 64mT contrast characteristics while improving the visibility of anatomical structures relevant for quantitative analysis.

The proposed pipeline incorporates automated localization, reorientation, image enhancement, tissue segmentation, and volumetric analysis within a single framework. A multi-channel 3D BasicUNet was trained on 75 paired neonatal 64mT/3T T2w datasets with 15 tissue classes. To preserve native tissue contrast, the 3T training images were locally harmonized to the corresponding 64mT scans using local histogram matching. The network simultaneously predicts enhanced images and tissue segmentations, while additional quality-control outputs provide information regarding segmentation confidence and enhancement consistency. The complete pipeline was implemented as a standalone Docker application with a runtime of less than five minutes per case.

Quantitative and qualitative evaluation on an independent cohort demonstrated that anatomy-aware enhancement improves visualization and delineation of cortical, deep gray matter, white matter, ventricular, and posterior fossa structures while maintaining native 64mT contrast relationships. Performance was highest for datasets with good native image quality and typical anatomy, whereas lower performance was primarily observed in cases with severe structural abnormalities or poor image quality that were underrepresented in the training cohort.

Importantly, to our knowledge, this is the first framework enabling automated multi-tissue neonatal brain segmentation and volumetric analysis directly from 64mT MRI. Evaluation on 40 paired control 64mT/3T datasets demonstrated excellent agreement between 64mT- and 3T-derived volumetric measurements across major tissue compartments. Although small systematic regional biases were observed, overall agreement remained high, supporting the feasibility of quantitative structural analysis at ultra-low field while also highlighting the need for dedicated 64mT normative references.

Overall, these findings support the integration of anatomy-aware enhancement into quantitative neonatal 64mT MRI workflows. Rather than replacing native 64mT imaging or attempting to reproduce conventional high-field appearance, the proposed framework aims to improve anatomical visibility while preserving native contrast information, thereby facilitating compatibility with existing neuroimaging tools and enabling future large-scale volumetric and population-level studies at ultra-low field.

## Limitations and future work

A key consideration when applying deep learning enhancement to 64mT MRI is the potential introduction of hallucinated or anatomically implausible image features. Although incorporation of anatomy-aware segmentation channel potentially partially reduces probability of generated implausible features, enhanced images should still always be interpreted alongside the native 64mT acquisition, particularly in regions associated with low QC confidence. While the proposed framework provides uncertainty and consistency estimates, future work should incorporate automated error detection, image quality assessment, and failure prediction.

Another limitation is the exclusion of motion-corrupted 64mT datasets since the current deep learning enhancement approach only amplifies artifacts rather then resolving them. Future versions could incorporate recently proposed alignedSENSE motion-correction reconstruction method for 64mT [27].

Furthermore, this study was performed on a relatively small paired cohort and should therefore be regarded as a feasibility study. Larger multi-centre datasets will be required to assess generalisability across different patient populations, developmental stages, and imaging conditions. Improved alignment of paired 64mT/3T datasets may further enhance performance. Clinical deployment will also require better representation of neonatal brain abnormalities, which are currently underrepresented in the training cohort. This limitation could be mitigated through synthetic 64mT data generation from existing 3T datasets [28] and integration with unsupervised anomaly detection frameworks [29].

Finally, extension of the framework to T1-weighted and diffusion MRI acquisitions [3], together with improved modelling of field-dependent tissue contrast, may facilitate broader translation of quantitative neonatal neuroimaging to ultra-low-field MRI. Further refinement of the segmentation component, including additional anatomical structures and improved cortical surface delineation [30, 31], could also enhance downstream morphometric analyses.

## Conclusion

We present a contrast-preserving, anatomy-aware framework for neonatal 64mT T2w brain MRI that jointly performs image enhancement and tissue segmentation. The proposed approach improves anatomical visibility, enables automated volumetric analysis, and shows strong agreement with corresponding 3T measurements. The standalone Docker application processes each case in under five minutes. These results support the potential of quantitative neonatal neuroimaging at low field and provide a foundation for future large-scale 64mT studies.

## Data Availability

The MRI datasets used in this study are not publicly available due to research ethics restrictions regarding patient confidentiality.

## Acknowledgments

We thank everyone involved in the acquisition and analysis of the datasets at the Department of Early Life Imaging at King’s College London and St Thomas’ Hospital. The authors thank all participants and their families.

This work was supported by Rosetrees Trust [2817415], the Bill and Melinda Gates Foundation (UNITY: Ultralow field Neuroimaging In The Young: INV-005798), the Wellcome/ EPSRC Centre for Medical Engineering at King’s College London [WT 203148/Z/16/Z], MRC Centre for Neurodevelopmental Disorders King’s College London [MR/N026063/1], MRC Senior Clinical Fellowship [MR/Y009665/1], the NIHR Clinical Research Facility (CRF) at Guy’s and St Thomas’ and by the National Institute for Health Research Biomedical Research Centre based at Guy’s and St Thomas’ NHS Foundation Trust and King’s College London. This study represents independent research in part funded by the National Institute for Health and Care Research (NIHR) Maudsley Biomedical Research Centre (BRC) at South London and Maudsley NHS Foundation Trust and King’s College London.

The views expressed are those of the authors and not necessarily those of the NHS, the NIHR or the Department of Health.

## Author contributions

P.C. and A.U. contributed equally to this work and are joint first authors. P.C. optimised the 64mT acquisition protocol, acquired datasets, performed analysis and evaluation. A.U. designed the contrast transfer pipeline, trained the networks and performed analysis. K.C., F.P, R.T. and I.T. contributed to optimization of the 64mT acquisition protocol and acquisition of the datasets. UNITY Consortium and S.C.R.W. provided the Hyperfine scanner. J.O.M., A.D.E., T.A. and J.V.H. provided the datasets and supervised various components of the project. M.R. provided the datasets, performed evaluation and supervised the project. All authors reviewed the manuscript.

